# Tuberculosis knowledge gaps and associated factors among suspects of tuberculosis in a peripheral emerging region of northwest Ethiopia: a case of Benishangul Gumuz

**DOI:** 10.1101/2022.07.11.22277486

**Authors:** Tekle Airgecho Lobie, Aklilu Abrham Roba

## Abstract

Good Knowledge about tuberculosis (TB) is a key element to have a favourable attitude and practice in the control of TB. However, in the resources limited peripheral emerging region of Ethiopia, there is a limited knowledge about it. Therefore, this study aimed to investigate the knowledge about TB symptoms, transmission, and prevention methods and identify the associated factors in Northwest Ethiopia.

**Methods:** A facility-based cross-sectional study was conducted among 391 TB suspects in two hospitals and seven health centres in the Benishangul Gumuz region. Composite scales were generated for knowledge about TB symptoms, route of transmission, and prevention methods. Data were analysed by Statistical Package for Social Sciences (SPSS) Version 25. Multivariate logistic regression was conducted to identify associated factors. A P-value less than 0.05 was considered statistically significant.

**Results:** From a total of 391 approached TB suspects, 383 (98%) participated in the study with the mean age of 33.34 ±14.10 years ranging from 11-89 years. There was very poor knowledge about TB symptoms (12%) but fair knowledge about TB transmission (61.6%), and prevention (57.4%). The previous history of treatment was significantly associated with the participants’ overall poor knowledge about TB symptoms (AOR 2.787, 95% CI=1.148-6.765), route of transmission (AOR=4.03, 95% CI=1.82-8.92), and prevention methods (AOR=4.89, 95% CI=2.18-10.99). Also, being illiterate was associated with poor knowledge about the route of TB transmission (AOR 11.39, 95% CI 2.15 - 60.33) prevention methods (AOR 13.28, 95% CI 2.69-65.73).

**Conclusion:** There was little knowledge about TB symptoms while fair knowledge about the mode of transmission and means of prevention in the Benishangul Gumuz region. Health education intervention particularly targeting TB symptoms, transmission, and prevention methods should be initiated through easily accessible media supported by effective strategies.

## Introduction

Tuberculosis (TB) is a communicable disease that is a major cause of ill health and one of the leading causes of death next to COVID-19 from a single infectious agent (Mycobacterium tuberculosis) [1]. Despite efforts being made to control TB, it remains a major global public health problem with 9.9 million fell ill with TB in 2020[1]. Africa bears 25% of all TB cases globally and Ethiopia is still among one of the 20 high TB and TB/HIV burden countries though the country managed to transition out of the list of the 30 high MDR-TB burden countries[2]. In 2018 the country has reported an estimated 165 000 TB cases, among which more than 30% (51 000) cases left not-notified or not diagnosed [3].

The most common clinical manifestation of TB is pulmonary distress which is slow and cleverly indirect at the onset. Patients typically have nonspecific complaints of malaise, fever, weight loss, cough, shortness of breath, chest pain, and night sweating. Sputum may be bloody if cavitary and purulent[4]. Treatment for new cases of drug-susceptible TB consists of a 6-month regimen of four first-line drugs: INH, RIF, EMB, and PZA. In order to avoid the occurrence of drug resistance, the drugs are provided in a combination of two or more [5]. In countries with a high TB burden, special emphasis has been given to the treatment of new TB cases by performing drug susceptibility tests that are relevant in the identification of resistant strains [6]. Treatment for MDR-TB is longer and more difficult to treat than drug-susceptible ones and threatens global progress towards the targets set by the End TB Strategy of the World Health Organization [7].

Inadequate knowledge about tuberculosis affects tuberculosis-related attitudes and practice of prevention which affects timely and appropriate treatment-seeking from health institutions[8]. Many studies in Ethiopia indicated that there was poor knowledge about the symptoms and cause of tuberculosis [9-13]. Similarly, studies indicate that the knowledge about TB transmission and prevention were either poor or inadequate[9, 11, 13]. A meta-analysis conducted in Ethiopia confirmed that poor knowledge about TB leads to a significant delay to seek treatment[14]. The treatment effectiveness is dependent on patients’ knowledge about the diseases, health service-seeking behaviour, and adherence to treatment [15, 16].

Globally, poverty and illiteracy were strongly associated with tuberculosis[17-21]. Poverty facilitates the transmission of TB, primarily through overcrowded and ventilated living or working conditions, high vulnerability due to poor nutrition and/or HIV, and delay in diagnosis [22, 23]. Financial limitations to the costs related to health services, transportation, accommodation, caterings, and loss of productivity affect TB prevention, and control efforts [24-26]. Moreover, in such communities, stigma and discrimination, rooted in misconceptions and utilization of the scarcely available health services contributed to poor TB treatment compliance and adherence. These factors contributed to advancing disease progression and further transmission [27-30].

The current study area is one of the resource-limited regions in Ethiopia where infrastructure and health facilities are very low. Literacy status in the study area was very low[31]. However, very little is known about TB-related knowledge in the area. Understanding the level of knowledge and the associated factors has paramount importance for the success of TB control programs. Therefore, this study aimed to assess knowledge about TB symptoms, transmission, and prevention methods with their associated factors in economically emerging and geographically peripheral region of Northwest Ethiopia.

## Materials and Methods

### Study area and design

An institution-based cross-sectional study was conducted in the two hospitals and seven health centers in the Benishangul Gumuz region namely Assosa general hospital, Pawe general hospital; Mambuk health center, Gilgelbeles health center, Felegeselem health center, Kemashi health center, and three health centers from Amhara National Regional State namely Dangila health center, Injibara health center, and Chagni health centers were included based on the presence of TB clinics and DOTS services. The details about the study area was mentioned somewhere[32].

### Sample size and population

The Sample size was calculated using the single population proportion formula (n = z^2^p (1-p)/d^2^, where n is the sample size, d is margin of error and p is the proportion of the event) based on multiple knowledge variables from prior studies. Also, G-power 3.1 [33] was used to determine the sample size for factors associated with TB knowledge and Attitude. The largest sample was considered (355). Thus, considering the 10% non-response rate, a total of 391 respondents were included in the study. All TB suspects referred to smear microscopy during nine months period were included regardless of their previous medical history. TB suspects less than 10 years of age were excluded.

### Data collection and tools

Data were collected through face-to-face interviews using a semi-structured questionnaire adapted from the WHO guide (WHO, 2008). Data collectors and supervisors were nurses who had been working in the TB clinic for at least 2 years at their respective health facilities. They received a 3-days training on the objectives of the study, data collection tool, interview protocol, and ethical aspects. Knowledge scores were generated for TB symptoms, route of transmission, and prevention methods using the mean of the number of correct answers by the study participants as a cutoff point to categorize good or poor knowledge score. The TB suspects who answered above the mean score were classified as having a good score, while those who scored below the mean were classified as having a poor knowledge score[13].

Dependent variables of the study were knowledge about TB symptoms, the route of transmission, prevention methods while the explanatory variables were: gender, age, educational and marital status, religion, residence, occupation, distance to health facility (on foot), income, family size, and history of TB treatment

### Statistical analysis

The collected data was entered in Epi-data version 3.1 and transferred and analyzed by Statistical Package for Social Sciences (SPSS) Version 25 (IBM Corp., Armonk, N.Y., USA)’. The normality of data was determined using Shapiro-Wilk’s test. All of the variables were normally distributed, Frequencies, proportions, and mean with standard deviations were used to describe the study population. Binary and multivariate logistic regression was done to extrapolate the presence and strength of association using crude and adjusted odds ratio (COR and AOR). A *P-value* less than 0.05 was considered statistically significant.

### Ethical considerations

The study was approved by the health research ethics review committee of Addis Ababa University, Faculty of Medicine, Department of Microbiology, Immunology, and Parasitology; and AHRI/ALERT research review committee with project Reg. No. PO12/13. Institutional support letters were obtained from regional, zonal, and local health administrations.

## Results

### Socio-demographic Characteristics

Of a total of 391 approached TB suspects, 383 (98%) participated in the study. The mean age of participants was 33.34 +14.10 years ranging from 11-89 years. The majority of respondents were male (60.3%), farmers (55.1%), rural residents (68.9%), Orthodox Christians (67.9%), and average monthly earnings of less than 500 Ethiopian Birr (51.2%). Also, one hundred sixty-five (43.1%) had never attended formal education. (Table-1)

**Table-1:** Socio-demographic characteristics of participants in Benishangul Gumuz

| Variables | Categories | Frequency (N=383) | Percent (%) |
| --- | --- | --- | --- |
| Gender | Male | 231 | 60.3 |
|  | Female | 152 | 39.7 |
| Occupation | Farmer | 211 | 55.1 |
|  | Merchant | 36 | 9.4 |
|  | Gov't employee | 40 | 10.4 |
|  | Student | 37 | 9.7 |
|  | Others* | 59 | 15.4 |
| Residence | Rural | 264 | 68.9 |
|  | Urban | 119 | 31.1 |
| Religion | Muslim | 88 | 23 |
|  | Orthodox | 260 | 67.9 |
|  | Protestant | 25 | 6.5 |
|  | Catholic | 7 | 1.8 |
|  | Others** | 3 | 0.8 |
| Educational status | No formal education | 165 | 43.1 |
|  | Grade 1-4 | 53 | 13.8 |
|  | Grade 5-8 | 79 | 20.6 |
|  | Grade 9-12 | 53 | 13.8 |
|  | Diploma | 25 | 6.5 |
|  | Bachelor and above | 8 | 2.1 |
| Income per annum<br>(Ethiopian Birr- ETB) | <500 | 196 | 51.2 |
|  | 500 | 29 | 7.6 |
|  | 501-1500 | 54 | 14.1 |
|  | 1501-3000 | 23 | 6 |
|  | 3001-5000 | 27 | 7 |
|  | >5000 | 54 | 14.1 |
| Distance to the nearest<br>health facility on foot | Less than an hour | 188 | 49.1 |
|  | One to Three hours | 59 | 15.4 |
|  | Three to Six hours | 3 | 0.8 |
|  | Half a day (12 hours?) | 114 | 29.8 |
|  | Two days | 2 | 0.5 |
|  | One day | 15 | 3.9 |
|  | One and half day | 2 | 0.5 |
| Marital status | Married | 211 | 55.1 |
|  | Single | 133 | 34.7 |
|  | Divorced | 26 | 6.8 |
|  | Widowed | 5 | 1.3 |
|  | Separated | 8 | 2.1 |
| Age | 11-20 | 66 | 17.2 |
|  | 21-30 | 146 | 38.1 |
|  | 31-40 | 83 | 21.7 |
|  | 41-50 | 49 | 12.8 |
|  | 51-60 | 24 | 6.3 |
|  | Above 60 | 15 | 3.9 |
| Ethnicity | Amhara | 237 | 61.9 |
|  | Gumuz | 31 | 8.1 |
|  | Agew | 31 | 8.1 |
|  | Oromo | 24 | 6.3 |
|  | Berta | 21 | 5.5 |
|  | Shinasha | 15 | 3.9 |
|  | Hadiya | 8 | 2.1 |
|  | Tigray | 5 | 1.3 |
|  | Others | 11 | 2.8 |

### Clinical characteristics

Most of the participants were presented with cough for more than two weeks (71.5 %), sputum production (30.5%), night sweating 144 (37.6%), fatigue 130 (33.9%), weight loss 96 (25.1%), fever 97 (25.3%) and difficulty to breath 82 (21.4%). Notably, fifty-six (14.6%) complained of combinations of all above mentioned clinical symptoms. (Fig-2)

**Fig 1:**
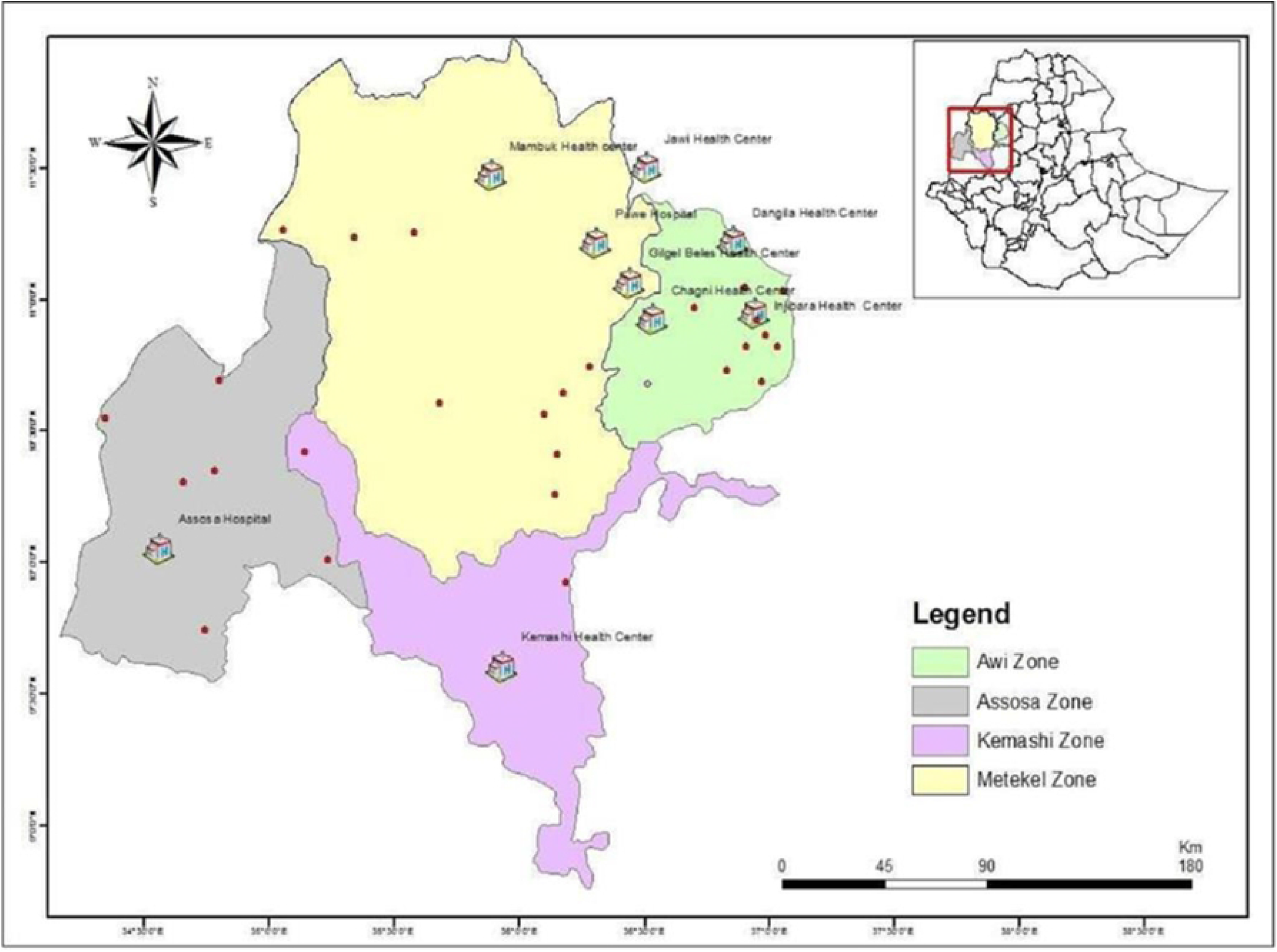
Map of the study area, Benishangul Gumuz region and surroundings.

**Figure 2:**
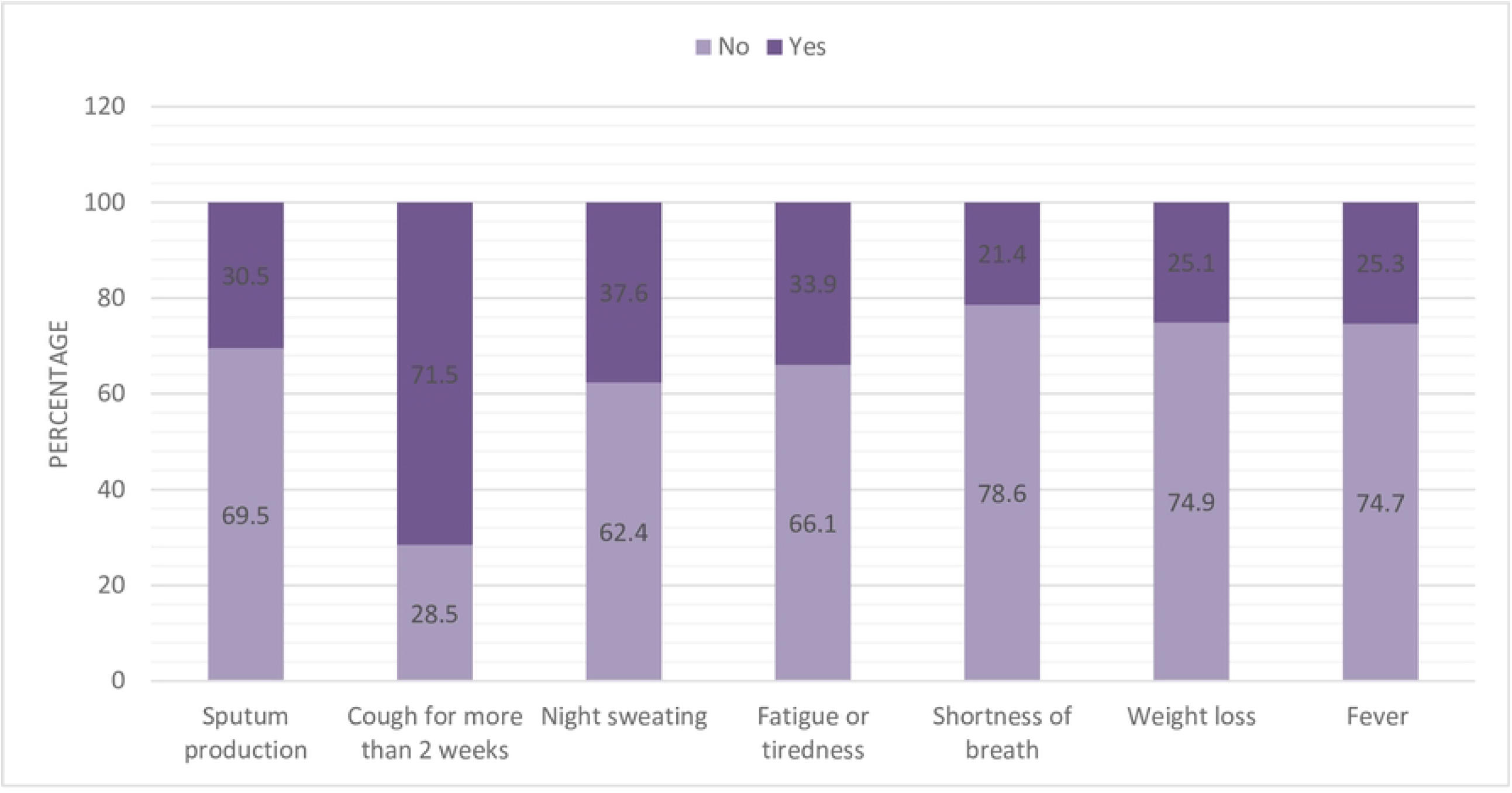
Clinical presentations of the study participants in Benishangul Gumuz

### Knowledge about TB symptoms, route of transmission, causes, and prevention

Regarding knowledge about the signs and symptoms of tuberculosis, participants correctly mentioned cough more than two weeks (54%), chest pain (24%), weakness or fatigue (26.4%), weight loss (17.8%), fever (13.6%). The majority (66.3%) correctly mentioned TB is transmitted through droplets during sneezing and coughing 58.2% mentioned covering the mouth and nose during coughing and sneezing was a possible means to prevent TB transmission. Remarkably high proportion (76.5 %) of the participants do not know the causative agent of TB. In general, only 12% of the respondents have good knowledge about TB symptoms, 61.6% have good knowledge about route of TB transmission, and 57.4% have good knowledge about TB prevention. (Table-2).

**Table 2:** Knowledge about TB symptoms, Route of transmission, Causes, and method of prevention in Benishangul Gumuz

| Variables | Responses | Frequency (N=383) | Percent (%) |
| --- | --- | --- | --- |
| TB Symptoms | Cough more than two weeks | 207 | 54 |
|  | Night Sweating | 87 | 22.7 |
|  | Chest pain | 92 | 24 |
|  | Shortness of breath | 74 | 19.3 |
|  | Fatigue or Weakness | 101 | 26.4 |
|  | Weight loss | 68 | 17.8 |
|  | Severe headache | 33 | 8.6 |
|  | Fever | 52 | 13.6 |
| Ways of transmission | Droplets during sneezing and coughing | 254 | 66.3 |
|  | Handshaking | 19 | 5 |
|  | Feeding together | 66 | 17.2 |
|  | Ingestion of raw milk | 56 | 14.6 |
|  | Curse | 2 | 0.5 |
| Cause of TB | Bacteria | 90 | 23.5 |
|  | Virus | 28 | 7.3 |
|  | Fungi | 1 | 0.3 |
|  | Curse | 2 | 0.5 |
|  | Cold temperature | 23 | 6 |
|  | Soil | 1 | 0.3 |
|  | Workload | 1 | 0.3 |
|  | I don't know | 235 | 61.4 |
|  | Others | 2 | 0.5 |
| Method of Prevention | Covering mouth/nose during coughing | 223 | 58.2 |
|  | Not shaking hands | 14 | 3.7 |
|  | Not eating together | 45 | 11.7 |
|  | Closing windows | 14 | 3.7 |
|  | Eating balanced diet | 34 | 8.9 |
|  | Praying | 8 | 2.1 |
|  | I don't know | 125 | 32.6 |
| Knowledge about symptoms of TB | Poor | 337 | 88 |
|  | Good | 46 | 12 |
| Knowledge about the transmission of TB | Poor | 147 | 38.4 |
|  | Good | 236 | 61.6 |
| Knowledge about the prevention of TB | Poor | 163 | 42.6 |
|  | Good | 220 | 57.4 |

The majority 324 (84.4%) of participants heard about TB from different sources majorly from health professionals and radio. Fig-3

**Fig 3:**
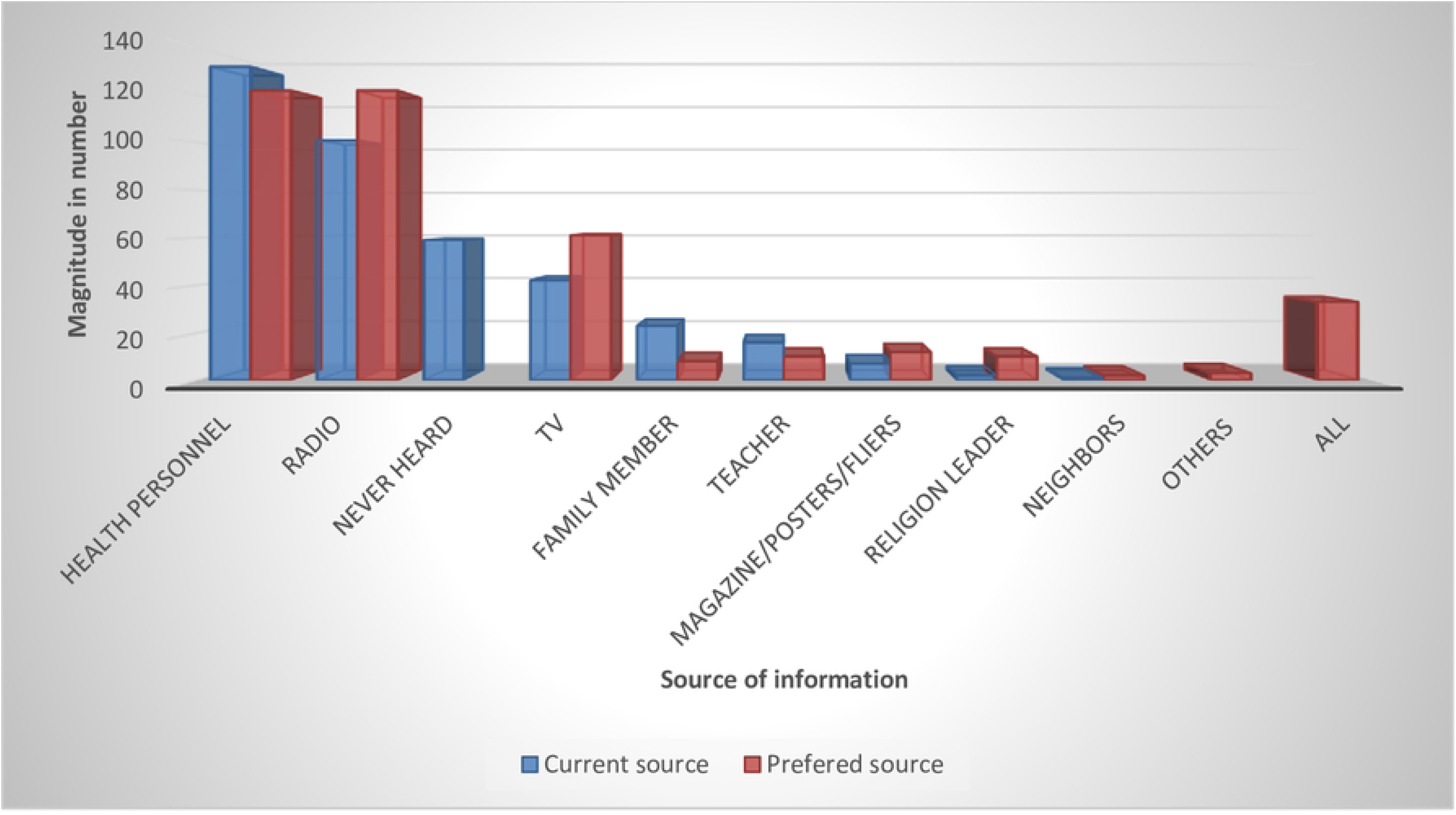
The current and preferred information sources about TB.

### The attitude of respondents towards TB

Most of the respondents feel TB is a severe (48%) and moderately severe (29.8%) disease in their locality. Around four in five respondents felt that TB affects everybody while 5.7% and 2.9% think the poor and PLWHA are affected, respectively. Asked what do they feel if they acquire TB, 32.4 %, 38.6%, and 14.1% expressed coping with it, fear, and sorrow, respectively. Also, 19.8% and 9.9% of participants mentioned long treatment duration and drug side-effects are the major critical issue. Only 47.2% would like to visit a health institution when they feel TB symptoms, with cost (16.4%) and distance (14.6%) being the most commonly presented reasons. The majority (94.8 %) knew TB could be cured and of them 92.7% mentioned anti-TB drugs (92.7 %). (Table-3)

**Table 3:**
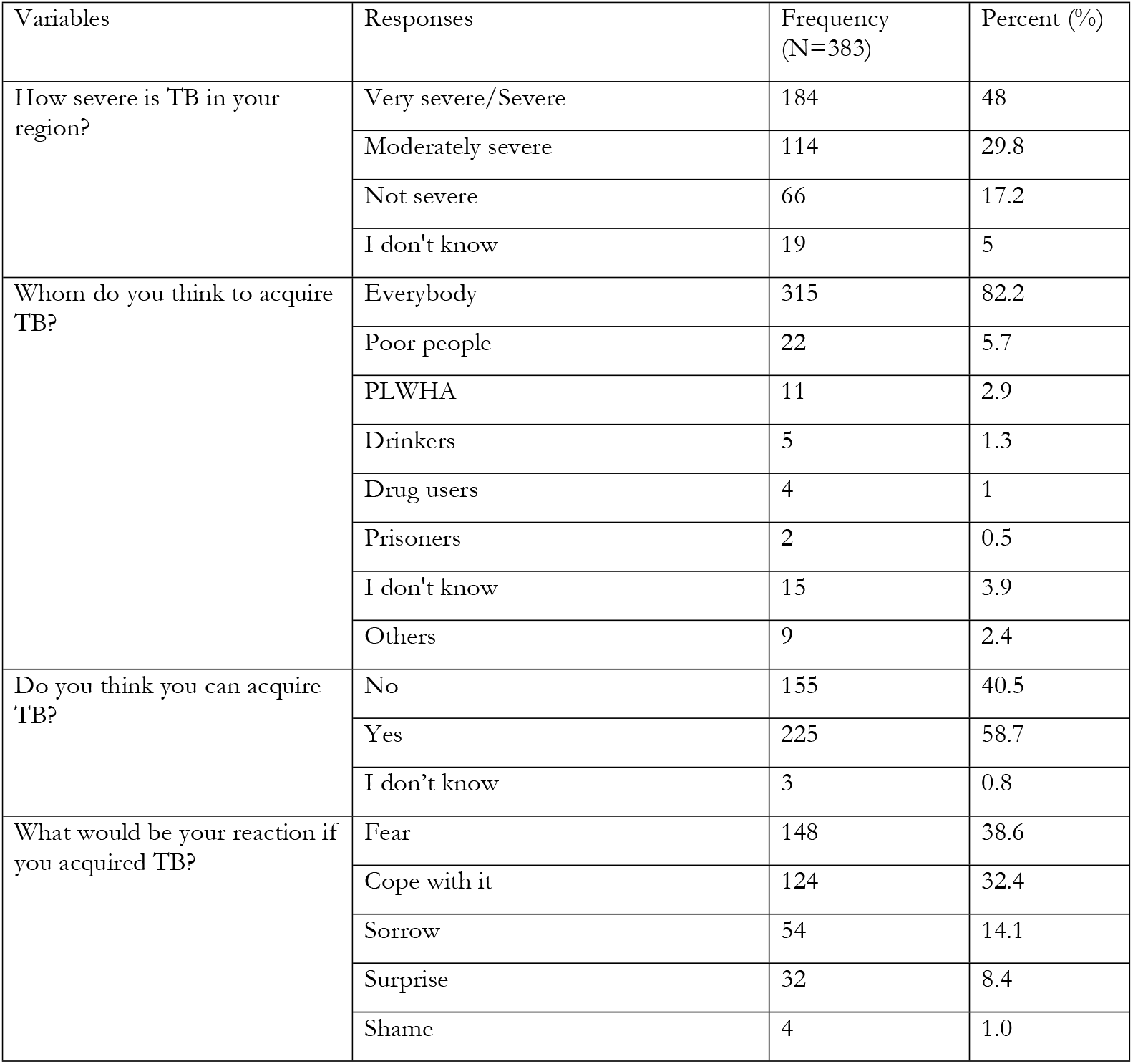
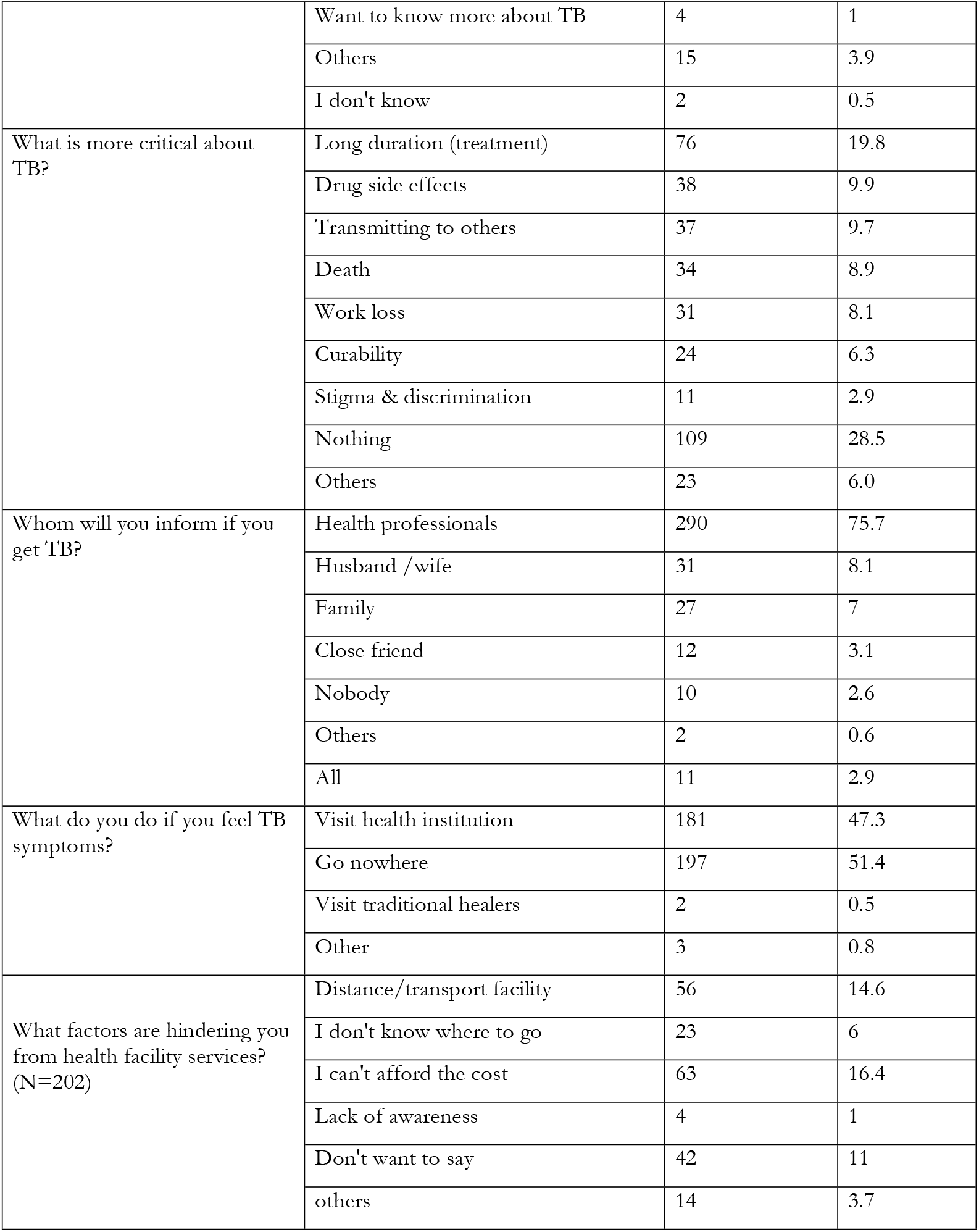
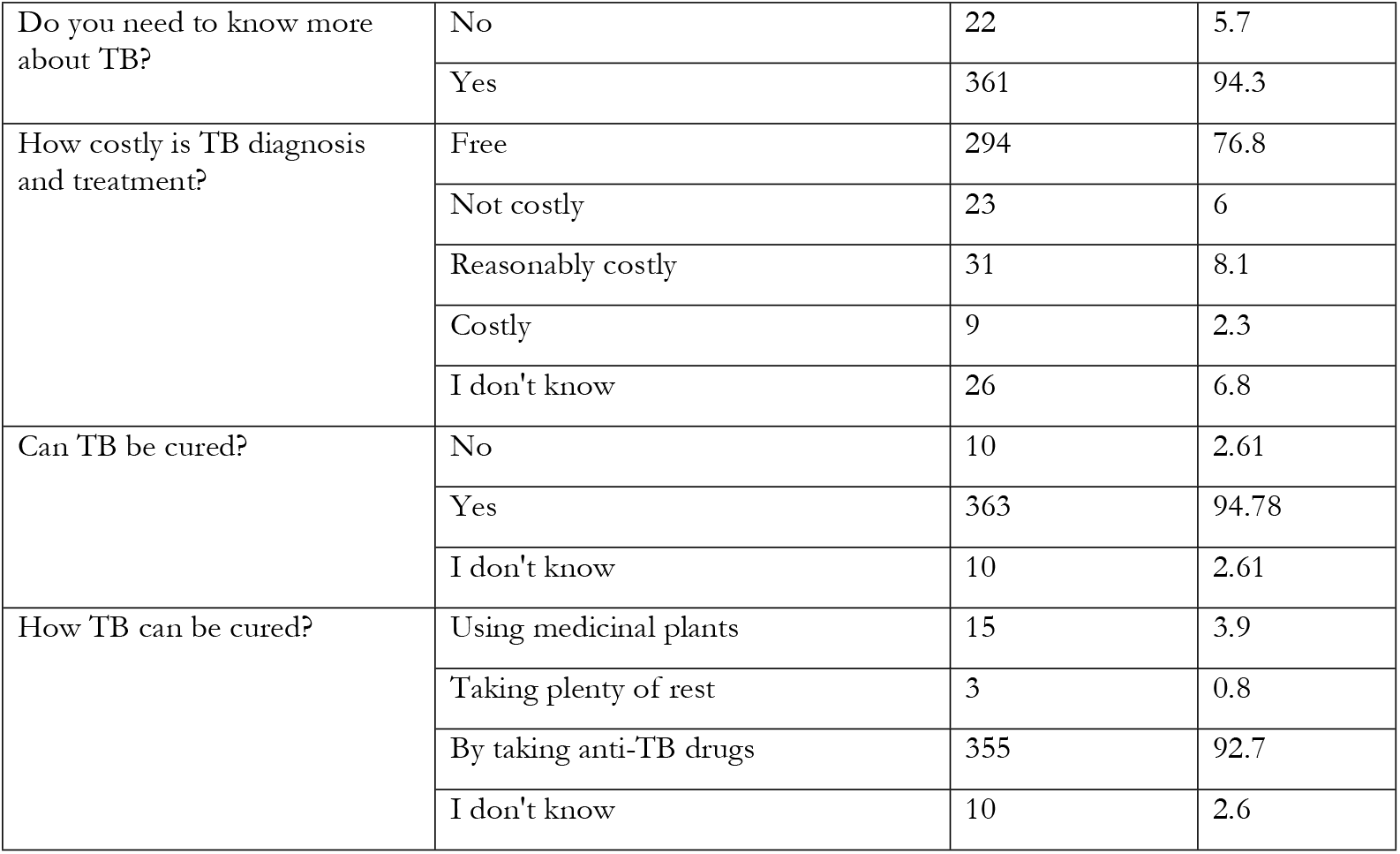
Attitude of study participants towards tuberculosis in Benishangul Gumuz

| Variables | Responses | Frequency<br>(N=383) | Percent (%) |
| --- | --- | --- | --- |
| How severe is TB in your region? | Very severe/Severe | 184 | 48 |
|  | Moderately severe | 114 | 29.8 |
|  | Not severe | 66 | 17.2 |
|  | I don't know | 19 | 5 |
| Whom do you think to acquire TB? | Everybody | 315 | 82.2 |
|  | Poor people | 22 | 5.7 |
|  | PLWHA | 11 | 2.9 |
|  | Drinkers | 5 | 1.3 |
|  | Drug users | 4 | 1 |
|  | Prisoners | 2 | 0.5 |
|  | I don't know | 15 | 3.9 |
|  | Others | 9 | 2.4 |
| Do you think you can acquire TB? | No | 155 | 40.5 |
|  | Yes | 225 | 58.7 |
|  | I don't know | 3 | 0.8 |
| What would be your reaction if you acquired TB? | Fear | 148 | 38.6 |
|  | Cope with it | 124 | 32.4 |
|  | Sorrow | 54 | 14.1 |
|  | Surprise | 32 | 8.4 |
|  | Shame | 4 | 1.0 |
|  | Want to know more about TB | 4 | 1 |
|  | Others | 15 | 3.9 |
|  | I don't know | 2 | 0.5 |
| What is more critical about TB? | Long duration (treatment) | 76 | 19.8 |
|  | Drug side effects | 38 | 9.9 |
|  | Transmitting to others | 37 | 9.7 |
|  | Death | 34 | 8.9 |
|  | Work loss | 31 | 8.1 |
|  | Curability | 24 | 6.3 |
|  | Stigma & discrimination | 11 | 2.9 |
|  | Nothing | 109 | 28.5 |
|  | Others | 23 | 6.0 |
| Whom will you inform if you get TB? | Health professionals | 290 | 75.7 |
|  | Husband /wife | 31 | 8.1 |
|  | Family | 27 | 7 |
|  | Close friend | 12 | 3.1 |
|  | Nobody | 10 | 2.6 |
|  | Others | 2 | 0.6 |
|  | All | 11 | 2.9 |
| What do you do if you feel TB symptoms? | Visit health institution | 181 | 47.3 |
|  | Go nowhere | 197 | 51.4 |
|  | Visit traditional healers | 2 | 0.5 |
|  | Other | 3 | 0.8 |
| What factors are hindering you from health facility services?<br>(N=202) | Distance/transport facility | 56 | 14.6 |
|  | I don't know where to go | 23 | 6 |
|  | I can't afford the cost | 63 | 16.4 |
|  | Lack of awareness | 4 | 1 |
|  | Don't want to say | 42 | 11 |
|  | others | 14 | 3.7 |
| Do you need to know more about TB? | No | 22 | 5.7 |
|  | Yes | 361 | 94.3 |
| How costly is TB diagnosis and treatment? | Free | 294 | 76.8 |
|  | Not costly | 23 | 6 |
|  | Reasonably costly | 31 | 8.1 |
|  | Costly | 9 | 2.3 |
|  | I don't know | 26 | 6.8 |
| Can TB be cured? | No | 10 | 2.61 |
|  | Yes | 363 | 94.78 |
|  | I don't know | 10 | 2.61 |
| How TB can be cured? | Using medicinal plants | 15 | 3.9 |
|  | Taking plenty of rest | 3 | 0.8 |
|  | By taking anti-TB drugs | 355 | 92.7 |
|  | I don't know | 10 | 2.6 |

### Factors associated with knowledge about the route of TB transmission, prevention methods, and symptom

The previous history of treatment was significantly associated with the participants’ overall knowledge about TB symptoms (AOR 2.787, 95% CI=1.148-6.765), transmission (AOR=4.03, 95% CI=1.82-8.92), and prevention methods (AOR=4.89, 95% CI=2.18-10.99).

Illiterate individuals and those with no TB treatment history were 11 times (AOR 11.39, 95% CI 2.15 - 60.33) and 4 times (AOR 4.03, 95% CI 1.82 - 8.92) more likely to have poor knowledge about the route of TB transmission compared to those with good knowledge, respectively. Table-4

**Table 4.** Factors associated with **knowledge about the route of transmission** related to tuberculosis.

| Factors | Categories | COR (95% CI) | AOR (95% CI) |
| --- | --- | --- | --- |
| Gender | Female | 1.49 (0.98 - 2.26) | 1.19 (0.71 - 2.00) |
|  | Male | 1 | 1 |
| Occupation | Farmers | 3.08 (1.35 - 7.06) * | 2.00 (0.56 - 7.14) |
|  | Merchant | 1.81 (0.64 - 5.16) | 1.84 (0.42 - 7.99) |
|  | Gov't employee | 0.77 (0.25 - 2.38) | 2.36 (0.44 - 12.63) |
|  | Others | 2.32 (0.90 - 5.94) | 2.29 (0.70 - 7.51) |
|  | Student | 1 | 1 |
| Residence | Urban | 0.51 (0.32 - 0.81) * | 0.74 (0.40 - 1.37) |
|  | Rural | 1 | 1 |
| Educational status | Illiterate | 7.06 (2.60 - 19.18) ** | 11.39 (2.15 - 60.33) * |
|  | Grade 1-4 | 2.21 (0.72 - 6.80) | 3.05 (0.56 - 16.61) |
|  | Grade 5-8 | 2.44 (0.84 - 7.09) | 2.55 (0.52 - 12.52) |
|  | Grade 9-12 | 1.47 (0.46 - 4.68) | 1.57 (0.34 - 7.22) |
|  | Higher education | 1 | 1 |
| Income status<br>(Ethiopian Birr) | <500 | 2.68 (1.33 - 5.40) * | 1.91 (0.85 - 4.31) |
|  | 500 | 2.84 (1.08 - 7.53) * | 2.74 (0.89 - 8.42) |
|  | 501-1500 | 1.90 (0.81 - 4.45) | 1.73 (0.65 - 4.58) |
|  | 1501-3000 | 2.69 (0.95 - 7.65) | 2.60 (0.78 - 8.72) |
|  | 3001-5000 | 1.47 (0.52 - 4.19) | 1.05 (0.32 - 3.45) |
|  | > 5000 | 1 | 1 |
| Distance | 3-6 hours | 0.96 (0.50 - 1.82) | 0.87 (0.40 - 1.89) |
|  | 1-2 days | 1.91 (0.69 - 5.33) | 1.65 (0.48 - 5.74) |
|  | < an hour | 0.91 (0.50 - 1.66) | 0.95 (0.45 - 1.99) |
|  | 1-3 hours | 1 | 1 |
| Marital status | Married | 1.05(0.17 - 6.41) | 0.48 (0.065 - 3.565) |
|  | Unmarried | 1.25 (0.20 - 7.74) | 0.56 (0.07 - 4.30) |
|  | Divorced | 0.57 (0.08 - 4.10) | 0.83 (0.10 - 6.79) |
|  | Widowed | 1 | 1 |
| Age group | 11-20 | 0.47 (0.15 - 1.45) | 1.38 (0.30 - 6.27) |
|  | 21-30 | 0.50(0.17 - 1.45) | 0.89 (0.24 - 3.34) |
|  | 31-40 | 0.67 (0.22 - 2.02) | 0.72 (0.19 - 2.71) |
|  | 41-50 | 0.47 (0.14 - 1.50) | 0.43 (0.11 - 1.68) |
|  | 51-60 | 0.63 (0.17 - 2.29) | 0.66 (0.15 - 2.98) |
|  | Above 60 | 1 | 1 |
| Family size | 1 | 0.90 (0.15 - 5.37) | 0.89 (0.12 - 6.80) |
|  | 2-4 | 1.31 (0.23 - 7.38) | 1.35 (0.19 - 9.50) |
|  | 5-7 | 1.18 (0.21 - 6.66) | 1.10 (0.15 - 7.85) |
|  | 8-10 | 2.13 (0.34 - 13.40) | 1.54 (0.19 - 12.22) |
|  | >10 | 1 | 1 |
| History of TB treatment | No | 2.68 (1.33 - 5.39) * | 4.03 (1.82 - 8.92) ** |
|  | Yes | 1 | 1 |

\*\* p-value less than 0.001, \* p-value less than 0.05; The reference category is 'Good' (N=383)

Illiterate individuals were 13 times (AOR 13.28, 95% CI 2.69-65.73), participants who earn less than 500 ETB were twice (AOR 2.40, 95% CI 1.077 - 5.335) and those with no TB treatment history were 4 times (AOR 4.89, 95% CI, 2.178-10.994) more likely to have poor knowledge about TB prevention methods compared to those with good knowledge. Table-5

**Table 5.**
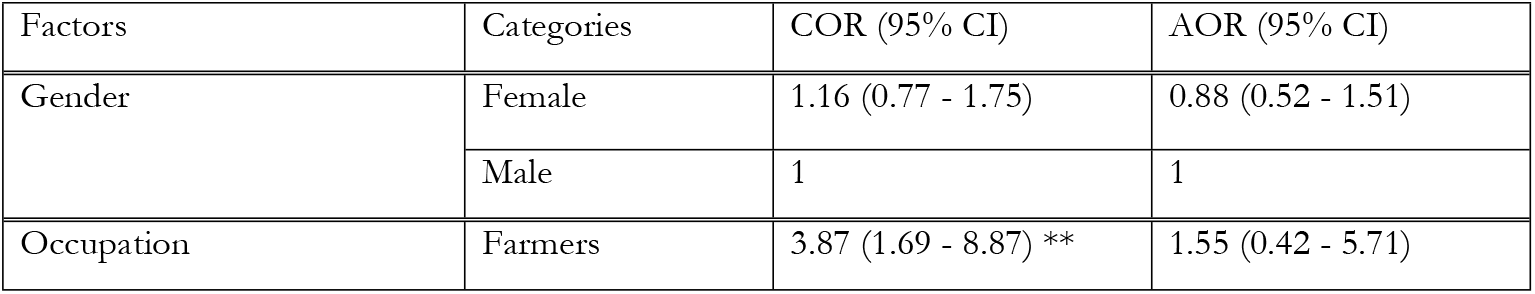
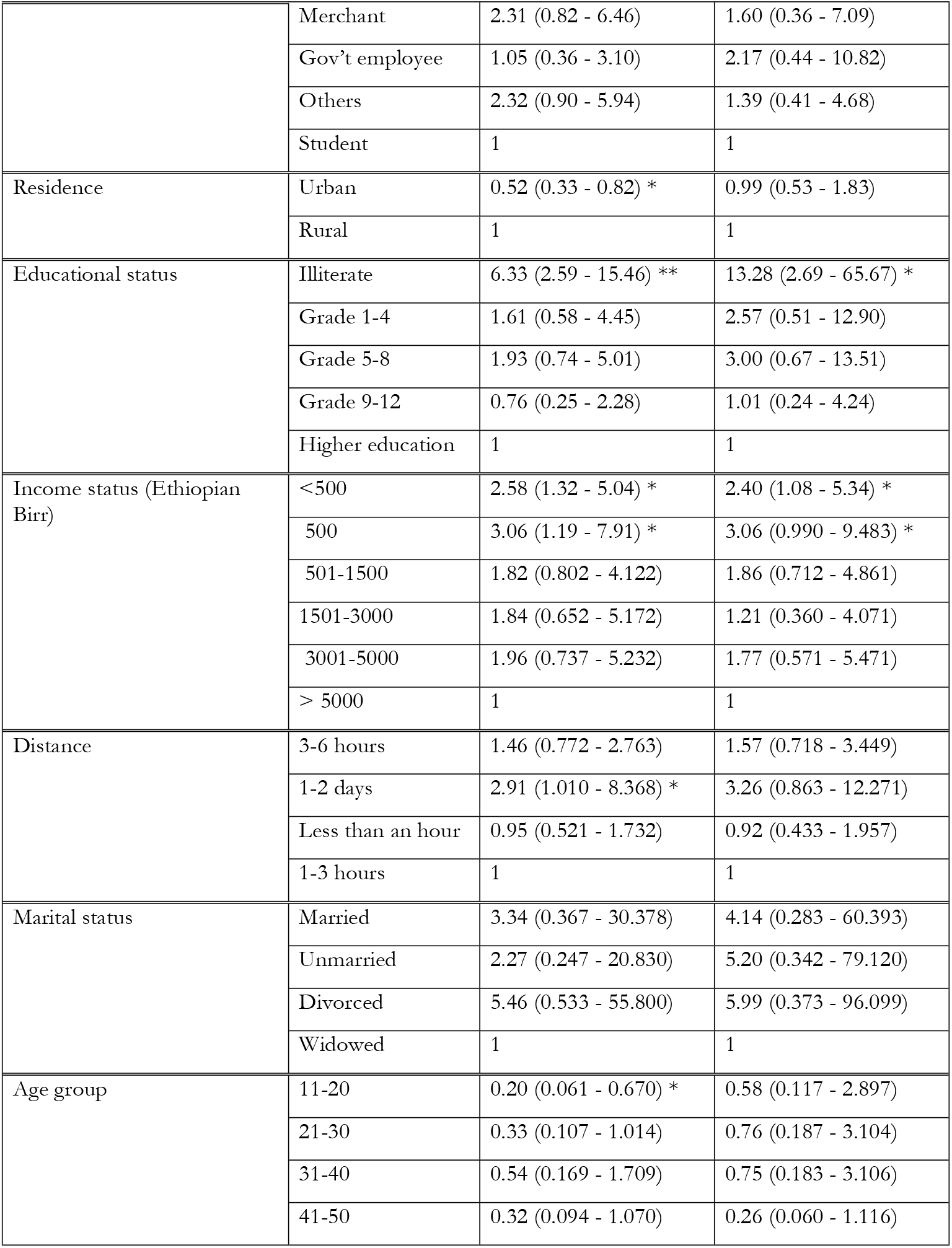
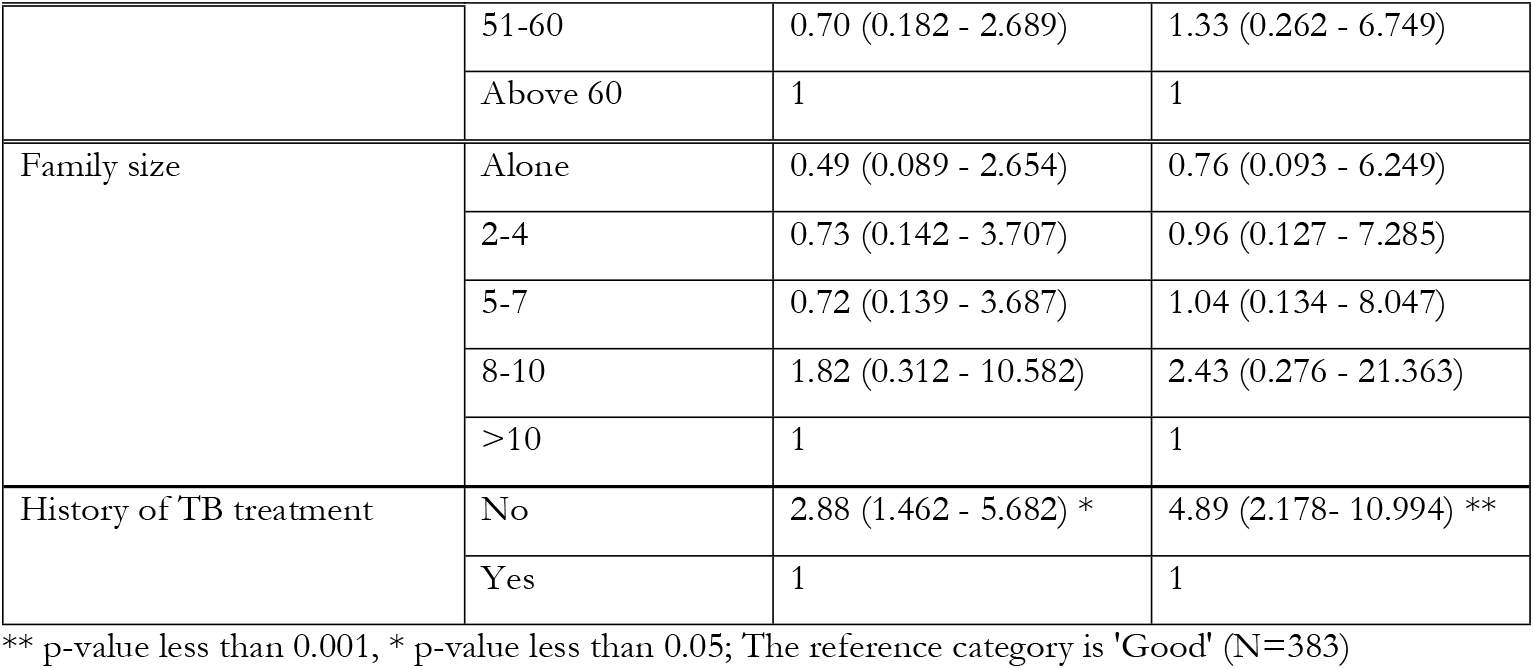
Factors associated with **knowledge about TB prevention in Benishangul Gumuz**

Individuals with no TB treatment history were three times (AOR 2.787, 95% CI 1.148 - 6.765) more likely to have poor knowledge about TB symptoms compared to those with good knowledge, respectively. Table-6

**Table-6:**
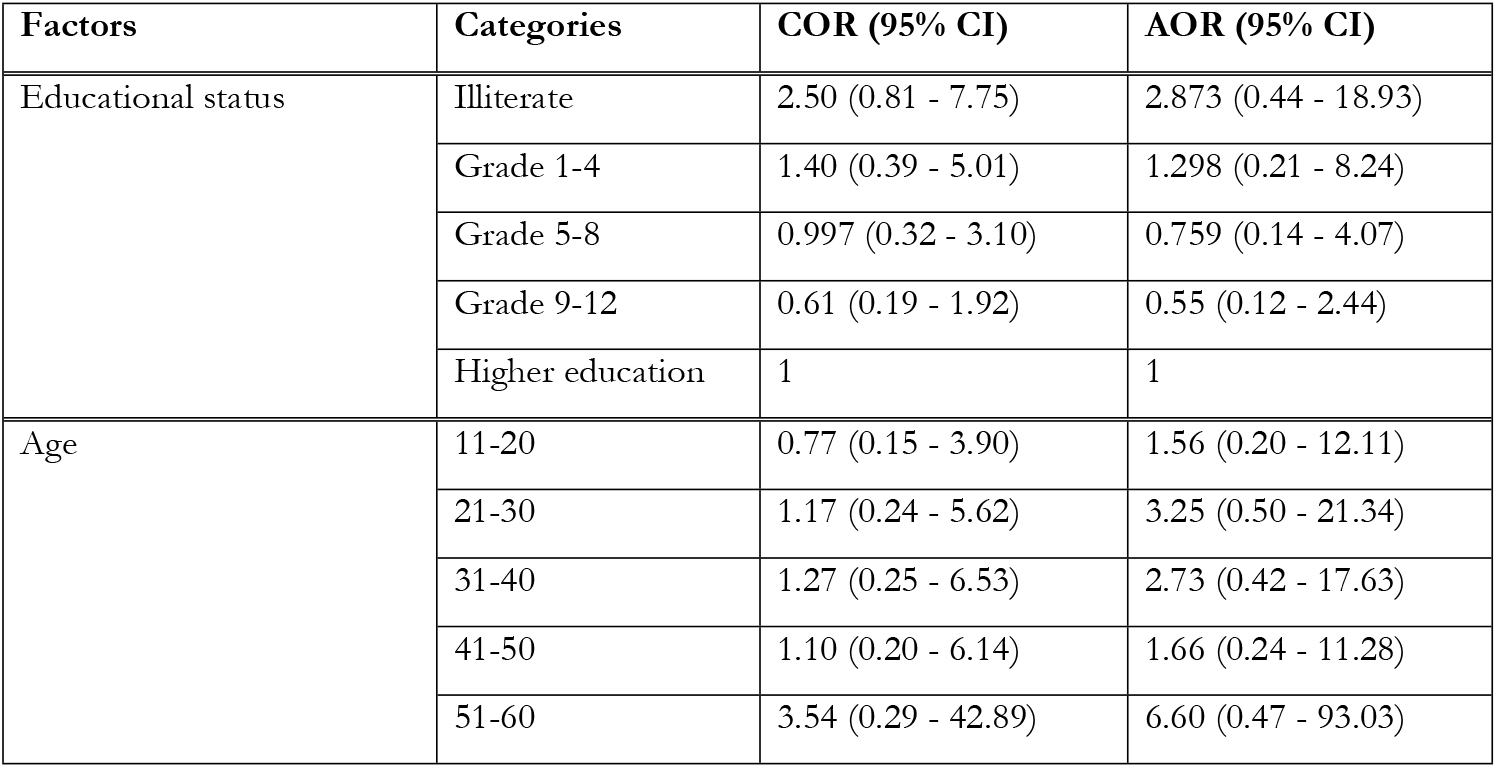
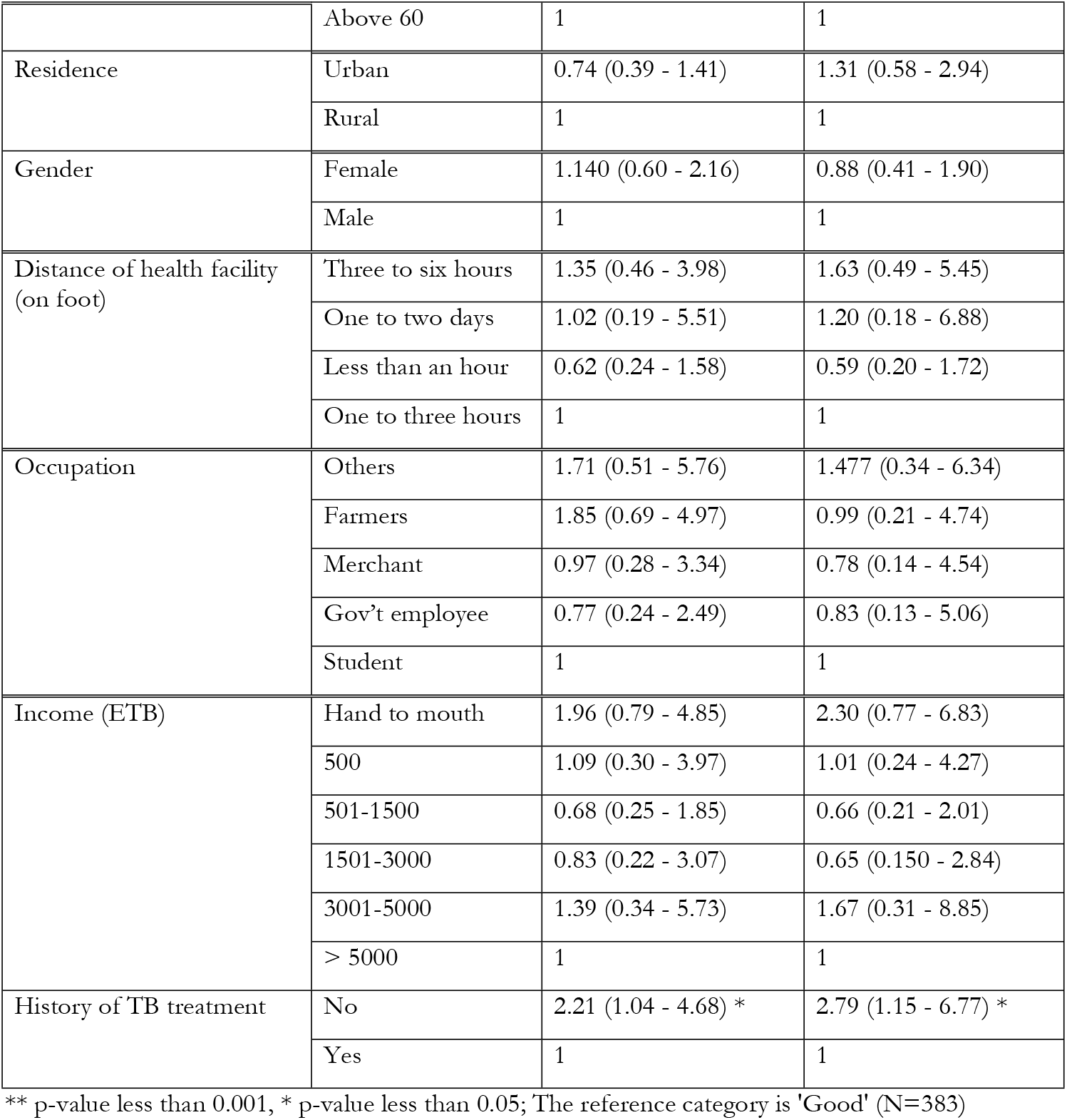
Factors associated with **knowledge about TB symptoms** related to tuberculosis in Benishangul Gumuz

| Factors | Categories | COR (95% CI) | AOR (95% CI) |
| --- | --- | --- | --- |
| Educational status | Illiterate | 2.50 (0.81 - 7.75) | 2.873 (0.44 - 18.93) |
|  | Grade 1-4 | 1.40 (0.39 - 5.01) | 1.298 (0.21 - 8.24) |
|  | Grade 5-8 | 0.997 (0.32 - 3.10) | 0.759 (0.14 - 4.07) |
|  | Grade 9-12 | 0.61 (0.19 - 1.92) | 0.55 (0.12 - 2.44) |
|  | Higher education | 1 | 1 |
| Age | 11-20 | 0.77 (0.15 - 3.90) | 1.56 (0.20 - 12.11) |
|  | 21-30 | 1.17 (0.24 - 5.62) | 3.25 (0.50 - 21.34) |
|  | 31-40 | 1.27 (0.25 - 6.53) | 2.73 (0.42 - 17.63) |
|  | 41-50 | 1.10 (0.20 - 6.14) | 1.66 (0.24 - 11.28) |
|  | 51-60 | 3.54 (0.29 - 42.89) | 6.60 (0.47 - 93.03) |
|  | Above 60 | 1 | 1 |
| Residence | Urban | 0.74 (0.39 - 1.41) | 1.31 (0.58 - 2.94) |
|  | Rural | 1 | 1 |
| Gender | Female | 1.140 (0.60 - 2.16) | 0.88 (0.41 - 1.90) |
|  | Male | 1 | 1 |
| Distance of health facility (on foot) | Three to six hours | 1.35 (0.46 - 3.98) | 1.63 (0.49 - 5.45) |
|  | One to two days | 1.02 (0.19 - 5.51) | 1.20 (0.18 - 6.88) |
|  | Less than an hour | 0.62 (0.24 - 1.58) | 0.59 (0.20 - 1.72) |
|  | One to three hours | 1 | 1 |
| Occupation | Others | 1.71 (0.51 - 5.76) | 1.477 (0.34 - 6.34) |
|  | Farmers | 1.85 (0.69 - 4.97) | 0.99 (0.21 - 4.74) |
|  | Merchant | 0.97 (0.28 - 3.34) | 0.78 (0.14 - 4.54) |
|  | Gov't employee | 0.77 (0.24 - 2.49) | 0.83 (0.13 - 5.06) |
|  | Student | 1 | 1 |
| Income (ETB) | Hand to mouth | 1.96 (0.79 - 4.85) | 2.30 (0.77 - 6.83) |
|  | 500 | 1.09 (0.30 - 3.97) | 1.01 (0.24 - 4.27) |
|  | 501-1500 | 0.68 (0.25 - 1.85) | 0.66 (0.21 - 2.01) |
|  | 1501-3000 | 0.83 (0.22 - 3.07) | 0.65 (0.150 - 2.84) |
|  | 3001-5000 | 1.39 (0.34 - 5.73) | 1.67 (0.31 - 8.85) |
|  | > 5000 | 1 | 1 |
| History of TB treatment | No | 2.21 (1.04 - 4.68) * | 2.79 (1.15 - 6.77) * |
|  | Yes | 1 | 1 |

\*\* p-value less than 0.001, \* p-value less than 0.05; The reference category is 'Good' (N=383)

## Discussion

This study found very poor knowledge about TB symptoms (12%) but fair knowledge about TB transmission (61.6%), and prevention (57.4%). Tuberculosis is regarded as a severe disease by 48% of respondents in their locality. However, only 47.3% reported to visit health institution when they feel they have TB symptoms. The previous history of treatment, educational status, and average monthly income shown varying degree of associations with tuberculosis knowledge components.

One of the fascinating findings of the study participants was almost all of them (94.78 %) reported TB could be cured by taking anti-TB drugs. This is comparable with 85.3% of a cross-sectional survey in seven regions and two city administrations of Ethiopia[34]. This information was disseminated majorly by health professionals (34.2%) and public radio (26.1%). This could be linked to the fact that most rural communities have better access to radio and health extension workers (community health workers) that live and work in their community. Similarly, studies from others areas of the country on the sources of information about TB reported a comparable finding [34, 35]. Therefore, program planners, researchers, and interventionists should consider radio and health professionals as a means for information dissemination.

Bacteria was mentioned as a cause of TB by 23.5% of participants while a large majority, 61.4%, reported they don’t know the cause. The knowledge about the cause of tuberculosis in the Benishangul-Gumuz region is as low as other Ethiopian studies: 37.7 % among prisoners in northern Ethiopia (37.7%)[36], 9.9 % of Tepi, South West Ethiopia [9], and 1.6% in Harar, Dire Dawa, and Jigjiga, Eastern Ethiopia[37], 25.8% of a nationwide cross-sectional TB KAP survey in Ethiopia[34], and 24.3% in Harar town[38]. Misconceptions about the cause of tuberculosis affect health-seeking behavior and utilization of freely provided TB services, thereby contributing to the dissemination of TB within the community [13]. The knowledge gap about the cause of TB may originate from overlooking the topic by health professionals.

Knowledge about TB transmission through droplets during sneezing and coughing was not adequate (only 67.3 %) and needs attention. Though there are comparable studies throughout the country, it could pause prevention and control efforts and thereby sustain the chain of TB transmission in the community [38-40]. Different studies have proven that coughing, sneezing, talking and also singing can transmit TB if appropriately mouths and noses were not covered[41-45]. This also agrees with a poor overall TB symptom-related knowledge (88%) in this study area. This finding agrees with similar studies conducted in Ethiopia [39, 46, 47]. Community health extension workers and health professionals should target this during health education session.

Knowledge about TB transmission was also reflected on TB prevention methods. Only 58.2% responded TB transmission can be prevented by covering the mouth and nose while coughing and sneezing with 42.6% participants had poor knowledge. A similar study elsewhere in the country revealed that 30.4% of participants do not know about the prevention and control of TB[47]. The wider gap seen in this regard could urge the need for additional strategies to be put in place to halt the continuing spread of TB burden in the peripheral regions through the provision of health education.

Although the majority (>80%) of respondents knew that everybody is at risk of acquiring TB, and believed that it is curable by taking anti-TB drugs from health institutions, 23.2% did not know that anti-tuberculosis drugs are being given freely. This finding agrees with finding from similar study [39] and adds the need for comprehensive health education addressing the available health services and related costs.

The current study identified a significant number of participants with unfavorable attitudes towards the severity of the disease, the population it affects, the possibility to be infected with it, reaction if they get the infection, and the action they would take. This is in agreement with other Ethiopian studies conducted elsewhere [9, 13, 48, 49]. TB program managers, administrators at health centers, and hospitals should strengthen the routine morning health education programs for all TB suspects and their care-takers.

One in three participant (32.4%) reported that they could cope with TB if they acquired it. This aligns with another similar study in Ethiopia[34]. However, more than half of them has fear (38.6%) and sorrow (14.1%). Attitudinal misconceptions also extend to fear of death (8.9%) and stigmatization and discrimination (2.9%). This indicates that some of the TB related knowledge were not translated to change their attitude and needs further sustained interventions.

To the best of our knowledge, this is the first study in the area to assess TB symptoms, route of transmission, and prevention methods and attitudes in the Benishangul-Gumuz region. This will help to strengthen TB control activities and support the scientific community, national and international policymakers to design appropriate interventions in the region. However, this study is not without limitations. As the study was conducted among the TB suspects, the knowledge and attitude may vary from the general community. Furthermore, the attitude component would provide deeper insight if it was supported by a qualitative study.

## Conclusions and Recommendations

There was little knowledge about TB symptoms while fair knowledge about the mode of transmission and means of prevention in the Benishangul Gumuz region. Moreover, substantial numbers of participants were not aware of the free service available. Health education intervention particularly targeting TB symptoms, transmission, and prevention methods should be initiated through easily accessible media supported by effective strategies.

## Data Availability

Data will be available with reasonable request through a corresponding author

## Authors’ contributions

**TA** participated in the conception, design and conduct of the study, data collection, data analysis, and drafting of the manuscript

**AA** conducted statistical analysis, interpretation of the findings, drafting the manuscript and revising it critically

## Acknowledgements

We are very grateful to the Armauer Hansen Research Institute (AHRI) for provision of funding and vehicle during the field work for data collection and Mr. Legesse Negash (Biostatistician at AHRI) for his constant assistance and guidance in statistical analysis.

## References

1. WHO. Global tuberculosis report 2021. Geneva: World Health Organization; 2021. Licence: CC BY-NC-SA 3.0 IGO. 2021.

2. World Health Organization. WHO global lists of high burden countries for tuberculosis (TB), TB/HIV and multidrug/rifampicin-resistant TB (MDR/RR-TB), 2021–2025: background document. 2021.

3. (WHO) WHO. Global Tuberculosis Report 2019. PubMed PMID: https://apps.who.int › iris › bitstream › handle › 9789241565714-eng › ua=1.

4. Fauci AS. Harrison’s principles of internal medicine: McGraw-Hill Education; 2015.

5. Sharma S, Mohan A. Extrapulmonary tuberculosis. Indian Journal of Medical Research. 2004;120(4):316.

6. Nachega JB, Chaisson RE. Tuberculosis drug resistance: a global threat. Clinical Infectious Diseases. 2003;36(Supplement_1):S24–S30.

7. Organization WH. WHO consolidated guidelines on tuberculosis: module 4: treatment: drug-resistant tuberculosis treatment: online annexes. 2020.

8. Sima BT, Belachew T, Abebe F. Knowledge, attitude and perceived stigma towards tuberculosis among pastoralists; Do they differ from sedentary communities? A comparative cross-sectional study. Plos One. 2017;12(7):17. doi: 10.1371/journal.pone.0181032. PubMed PMID: WOS:000443975500023.

9. Angelo AT, Geltore TE, Asega T. Knowledge, Attitude, and Practices Towards Tuberculosis Among Clients Visiting Tepi General Hospital Outpatient Departments, 2019. Infection and Drug Resistance. 2020;13:4559–68. doi: 10.2147/idr.s287288. PubMed PMID: WOS:000600626000002.

10. Legesse M, Ameni G, Mamo G, Medhin G, Shawel D, Bjune G, et al. Knowledge and perception of pulmonary tuberculosis in pastoral communities in the middle and Lower Awash Valley of Afar region, Ethiopia. Bmc Public Health. 2010;10:11. doi: 10.1186/1471-2458-10-187. PubMed PMID: WOS:000277609900001.

11. Bati J, Legesse M, Medhin G. Community’s knowledge, attitudes and practices about tuberculosis in Itang Special District, Gambella Region, South Western Ethiopia. Bmc Public Health. 2013;13:9. doi: 10.1186/1471-2458-13-734. PubMed PMID: WOS:000323117300001.

12. Tolossa D, Medhin G, Legesse M. Community knowledge, attitude, and practices towards tuberculosis in Shinile town, Somali regional state, eastern Ethiopia: a cross-sectional study. Bmc Public Health. 2014;14:13. doi: 10.1186/1471-2458-14-804. PubMed PMID: WOS:000340802700001.

13. Datiko DG, Habte D, Jerene D, Suarez P. Knowledge, attitudes, and practices related to TB among the general population of Ethiopia: Findings from a national cross-sectional survey. Plos One. 2019;14(10):16. doi: 10.1371/journal.pone.0224196. PubMed PMID: WOS:000532647400013.

14. Alene M, Assemie MA, Yismaw L, Gedif G, Ketema DB, Gietaneh W, et al. Patient delay in the diagnosis of tuberculosis in Ethiopia: a systematic review and meta-analysis. BMC Infectious Diseases. 2020;20(1). doi: 10.1186/s12879-020-05524-3.

15. Lawn S, Afful B, Acheampong J. Pulmonary tuberculosis: diagnostic delay in Ghanaian adults. The International Journal of Tuberculosis and Lung Disease. 1998;2(8):635–40.

16. Demissie M, Lindtjorn B, Berhane Y. Patient and health service delay in the diagnosis of pulmonary tuberculosis in Ethiopia. BMC Public Health. 2002;2(1):23.

17. Muniyandi M, Ramachandran R, Gopi P, Chandrasekaran V, Subramani R, Sadacharam K, et al. The prevalence of tuberculosis in different economic strata: a community survey from South India. The International Journal of Tuberculosis and Lung Disease. 2007;11(9):1042–5.

18. Barter DM, Agboola SO, Murray MB, Bärnighausen T. Tuberculosis and poverty: the contribution of patient costs in sub-Saharan Africa–a systematic review. BMC public health. 2012;12(1):1–21.

19. Oxlade O, Murray M. Tuberculosis and poverty: why are the poor at greater risk in India? PloS one. 2012;7(11):e47533.

20. Pathak D, Vasishtha G, Mohanty SK. Association of multidimensional poverty and tuberculosis in India. Bmc Public Health. 2021;21(1):12. doi: 10.1186/s12889-021-12149-x. PubMed PMID: WOS:000717504400001.

21. Erlinger S, Stracker N, Hanrahan C, Nonyane BAS, Mmolawa L, Tampi R, et al. Tuberculosis patients with higher levels of poverty face equal or greater costs of illness. International Journal of Tuberculosis and Lung Disease. 2019;23(11):1205–12. doi: 10.5588/ijtld.18.0814. PubMed PMID: WOS:000496915300012.

22. Marais B, Hesseling A, Cotton M. Poverty and tuberculosis: is it truly a simple inverse linear correlation? European Respiratory Journal. 2009;33(4):943–4.

23. Organization WH. Addressing poverty in TB control: options for national TB control programmes: World Health Organization; 2005.

24. Ahn DI, Organization WH. Addressing poverty in TB control: options for national TB control programmes. 2005.

25. Nhlema B, Kemp J, Steenbergen G, Theobald G, Tang S, Squire B. The state of existing knowledge about TB and poverty. Int J Tuberc Lung Dis. 2003;7(suppl 2):116.

26. Kamolratanakul P, Sawert H, Kongsin S, Lertmaharit S, Sriwongsa J, Na-Songkhla S, et al. Economic impact of tuberculosis at the household level. The International Journal of Tuberculosis and Lung Disease. 1999;3(7):596–602.

27. Yousif T, Mahmoud A, Khayat I. Survey of KAP: enhanced response to TB ACSM. Middle East J Family Med. 2009;7:7–13.

28. Irani L, Kabalimu T, Kasesela S. Knowledge and healthcare seeking behaviour of pulmonary tuberculosis patients attending Ilala District Hospital, Tanzania. Tanzania Journal of Health Research. 2008;9(3):169–73.

29. Khan JA, Irfan M, Zaki A, Beg M, Hussain SF, Rizvi N. Knowledge, attitude and misconceptions regarding tuberculosis in Pakistani patients. Journal of Pakistan Medical Association. 2006;56(5):211.

30. Lawn S, Shattock R, Griffin G. Delays in the diagnosis of tuberculosis: a great new cost [letter]. International Journal of Tuberculosis and Lung Disease. 1997;1(5):485–6.

31. MoE. ESAA 2010 E.C. (2017/18). 2018.

32. Lobie TA, Woldeamanuel Y, Asrat D, Beyene D, Bjørås M, Aseffa A. Genetic diversity and drug resistance pattern of Mycobacterium tuberculosis strains isolated from pulmonary tuberculosis patients in the Benishangul Gumuz region and its surroundings, Northwest Ethiopia. PloS one. 2020;15(4):e0231320.

33. Faul F, Erdfelder E, Buchner A, lang A. Statistical power analyses using G*Power 3.1: Tests for correlation and regression analyses. Behavior Research Methods, 41, 1149–1160. doi:10.3758/BRM.41.4.1149. 2009.

34. Datiko DG, Habte D, Jerene D, Suarez P. Knowledge, attitudes, and practices related to TB among the general population of Ethiopia: Findings from a national cross-sectional survey. PLoS ONE. 2019;14(10). doi: 10.1371/journal.pone.0224196.

35. Mesfin MM, Tasew TW, Tareke IG, Mulugeta GWM, Richard MJ. Community knowledge, attitudes and practices on pulmonary tuberculosis and their choice of treatment supervisor in Tigray, northern Ethiopia. Ethiopian Journal of Health Development. 2005;19(Special issue):21–7. PubMed PMID: CABI:20063001858.

36. Adane K, Spigt M, Johanna L, Noortje D, Abera SF, Dinant GJ. Tuberculosis knowledge, attitudes, and practices among northern Ethiopian prisoners: Implications for TB control efforts. Plos One. 2017;12(3):15. doi: 10.1371/journal.pone.0174692. PubMed PMID: WOS:000399174800073.

37. Abebe DS, Biffa D, Bjune G, Ameni G, Abebe F. Assessment of knowledge and practice about tuberculosis among eastern Ethiopian prisoners. International Journal of Tuberculosis and Lung Disease. 2011;15(2):228–33. PubMed PMID: WOS:000287011500016.

38. Seyoum A, Legesse M. Knowledge of tuberculosis (TB) and human immunodeficiency virus (HIV) and perception about provider initiated HIV testing and counselling among TB patients attending health facilities in Harar town, Eastern Ethiopia. Bmc Public Health. 2013;13. doi: 10.1186/1471-2458-13-124. PubMed PMID: WOS:000317762900001.

39. Esmael A, Ali I, Agonafir M, Desale A, Yaregal Z, Desta K. Assessment of patients’ knowledge, attitude, and practice regarding pulmonary tuberculosis in eastern Amhara regional state, Ethiopia: cross-sectional study. The American journal of tropical medicine and hygiene. 2013;88(4):785.

40. Mesfin MM, Tasew TW, Tareke IG, Mulugeta GW, Richard MJ. Community knowledge, attitudes and practices on pulmonary tuberculosis and their choice of treatment supervisor in Tigray, northern Ethiopia. Ethiopian Journal of Health Development. 2005;19(I):21.

41. Loudon RG, Spohn SK. Cough frequency and infectivity in patients with pulmonary tuberculosis. American Review of Respiratory Disease. 1969;99(1):109–11.

42. Duguid J. The size and the duration of air-carriage of respiratory droplets and droplet-nuclei. Epidemiology & Infection. 1946;44(6):471–9.

43. Loudon RG, Roberts RM. Singing and the dissemination of tuberculosis. American Review of Respiratory Disease. 1968;98(2):297–300.

44. Patterson B, Morrow C, Singh V, Moosa A, Gqada M, Woodward J, et al. Detection of Mycobacterium tuberculosis bacilli in bio-aerosols from untreated TB patients. Gates open research. 2017;1.

45. Turner R, Birring S, Darmalingam M, Hooper R, Kunst H, Matos S, et al. Daily cough frequency in tuberculosis and association with household infection. The International Journal of Tuberculosis and Lung Disease. 2018;22(8):863–70.

46. Bati J, Legesse M, Medhin G. Community’s knowledge, attitudes and practices about tuberculosis in Itang special district, Gambella region, south western Ethiopia. BMC public health. 2013;13(1):1–9.

47. Abebe D, Biffa D, Bjune G, Ameni G, Abebe F. Assessment of knowledge and practice about tuberculosis among eastern Ethiopian prisoners. The International journal of tuberculosis and lung disease. 2011;15(2):228–33.

48. Sima BT, Belachew T, Abebe F. Health care providers’ knowledge, attitude and perceived stigma regarding tuberculosis in a pastoralist community in Ethiopia: A cross-sectional study 11 Medical and Health Sciences 1117 Public Health and Health Services. BMC Health Services Research. 2019;19(1). doi: 10.1186/s12913-018-3815-1.

49. Bezawit Temesgen S, Tefera B, Fekadu A. Knowledge, attitude and perceived stigma towards tuberculosis among pastoralists; do they differ from sedentary communities? A comparative cross-sectional study. PLoS ONE. 2017;12(7):e0181032–e. doi: 10.1371/journal.pone.0181032. PubMed PMID: CABI:20173294671.

